# Divergent Causes of Death After Ischemic and Hemorrhagic Stroke in the Reperfusion Era: A Contemporary Cohort Study

**DOI:** 10.64898/2026.11.03.26367230

**Authors:** Vishnu Kadiyala, Alexis N. Simpkins, Harpriya Chugh, Ryan Czarny, Audrey Uy-Evanado, Nichole Bosson, Nicholas Tatonetti, Kyndaron Reinier, Sumeet S. Chugh

## Abstract

**Background:** Contemporary data on long-term, cause-specific mortality among stroke survivors are sparse, and much of the existing evidence derives from studies performed before the modern reperfusion era. We sought to characterize long-term cause-specific mortality among stroke survivors to discharge (differentiated by stroke subtype) when compared to in-hospital stroke decedents treated under contemporary standards of care.

**Methods:** Using a hybrid billing-code/imaging and large language model-assisted chart-review pipeline, we identified adults hospitalized for ischemic or hemorrhagic stroke in the Observational Study of Cardiac Arrest Risk (OSCAR) cohort (Cedars-Sinai Health System, Los Angeles) between 2017 and 2020, followed through June 30, 2025. Cause of death was determined and classified into 10 categories.

**Results:** Among 1,344 stroke patients (mean age 74.0 ± 14.9 years; ischemic n=1,036 [77.1%], hemorrhagic n=308 [22.9%]), 203 (15.1%) did not survive the index stroke. Of the 1,141 discharged survivors, 625 (54.8%) died over a median follow-up of 5.2 years. Cerebrovascular causes predominated during index stroke hospitalization (36.0%) but accounted for only 15.5% of post-stroke discharge deaths. Among ischemic stroke survivors, cardiac disease was the leading cause of death (31.6%), with sudden cardiac arrest alone accounting for 10.9% of deaths. Among hemorrhagic stroke survivors, cancer (38.9%) was the leading cause of death. Despite these divergent cause-of-death profiles, overall survival did not differ significantly by subtype (median 5.5 vs 5.8 years; *P*=0.90).

**Conclusions:** In this contemporary, reperfusion-era cohort, causes of death shifted from cerebrovascular predominance in the acute phase to heart disease/cardiac arrest after ischemic stroke, and malignancy after hemorrhagic stroke. These findings argue for development of subtype-specific post-stroke surveillance, including structured longitudinal cardiac and sudden-death risk assessment for ischemic stroke survivors.

## INTRODUCTION

Stroke is among the leading causes of death and disability worldwide. In 2019, this condition was the second-leading cause of death globally and accounted for more than 100 million prevalent cases, reflecting a large and expanding population of survivors^1^. For these survivors, a critical knowledge gap remains, particularly in the current reperfusion era: what causes longer-term mortality after stroke? Accurate characterization of these causes is an essential requirement for development of approaches for post-stroke surveillance in the reperfusion era.

Much of the prevailing understanding of mortality in stroke survivors comes from a small number of population-based cohorts assembled between the 1980s and the early 2000s: the Danish contribution to the WHO MONICA Project^2^, the Perth Community Stroke Study^3,4^, the Rochester, Minnesota, population^5^, and the Northern Manhattan Stroke Study^6^. Together these studies suggest that patterns of mortality after stroke are highly temporal. In the acute and subacute period, mortality is dominated by the index neurological injury and complications related to malignant brain edema and herniation. Thereafter, the cause-of-death profile shifts, with cardiovascular disease and recurrent stroke predominating among medium-term survivors and an increasing contribution from non-vascular causes over longer follow-up.

Three considerations limit how well this evidence applies to stroke patients treated today. First, these patterns have largely been described in ischemic stroke and may not extend to hemorrhagic stroke, which carries a far higher acute case fatality^7^ and a distinct long-term cause-of-death profile^8^. Pooling of these stroke subtypes may therefore obscure two clinically distinct mortality trajectories. Second, cardiac death, and specifically sudden or dysrhythmic death, after stroke is a plausible consequence of the bidirectional brain–heart relationship, whereby autonomic dysregulation, catecholamine surge, and neurogenic myocardial injury may precipitate cardiac events^9–11^; this constellation has been called the stroke–heart syndrome^12^. However, sudden and dysrhythmic death after stroke remains under-recognized and under-adjudicated in long-term cause-of-death series^11,13–15^. Third, nearly all these cohorts were assembled before, or at the dawn of, the modern reperfusion era. Even the more recent Oxford Vascular Study, which characterized the time course of cause-specific mortality in 1,606 ischemic stroke patients recruited between 2002 and 2014, predates routine mechanical thrombectomy as standard of care^16^. Intravenous thrombolysis was established after the pivotal 1995 trial of tissue plasminogen activator^17^, and endovascular thrombectomy did not become standard care for large-vessel occlusion until the five randomized trials of 2015 and the pooled HERMES analysis that followed^18–23^. Across this same interval, population-based registers have documented substantial declines in stroke case fatality^24^.

Contemporary cause-of-death data, by contrast, remain sparse and heavily weighted toward the short term, with most reports describing mortality only through the first days to weeks after the index event^25^. Far less is known about how stroke survivors die over the subsequent years, now that more patients survive the index event with preserved function. We therefore sought to characterize the causes of mortality among stroke survivors in the modern reperfusion era, with particular attention to differences by stroke subtype and discharge location, and compare these patterns to causes of death among patients who died during initial hospitalization for stroke.

## METHODS

### Study design

Detailed methods for the Observational Study of Cardiac Arrest Risk (OSCAR) study have been published previously^26^. Briefly, the OSCAR cohort is an electronic health records (EHR) study of Los Angeles County residents with at least one patient encounter per year in the Cedars-Sinai Health System during two consecutive years in the enrollment period from 2017 to 2020. Follow-up began on the date of the second qualifying visit (as early as Jan. 1, 2017 to as late as Dec. 31, 2020). EHR data were obtained from all outpatient and inpatient encounters across the health care system from year 2012 onwards, in order to establish the baseline EHR profile for all included patients.

For the current study, we included patients aged ≥35 years with a hospitalization for stroke after the start of follow-up until June 30, 2020, in OSCAR. Stroke patients were identified with a combined use of International Classification of Diseases, 10th Revision (ICD-10) codes and the ChartScope platform (described below).

Survival time was defined as the interval from the index stroke discharge date to death from any cause, or until the end of follow-up (June 30, 2025) for patients still alive. Baseline comorbidities were defined by ICD-10 diagnosis codes, supplemented by procedure data as appropriate as detailed in our group’s OSCAR design paper^26^.

This study is reported in accordance with the Strengthening the Reporting of Observational Studies in Epidemiology (STROBE) guideline^27^.

### Hybrid Billing Code/Imaging Code and LLM-based Approach for identification of stroke hospitalizations

#### Billing and imaging code-based algorithm for incident stroke

We identified patients admitted during the follow-up period (January 1, 2017 to June 30, 2020) with a diagnosis of stroke who had both 1) a final stroke-related diagnosis or billing ICD-10 code and 2) a procedure code indicating evidence of brain imaging during that admission, described below.

We classified stroke-related ICD-10 billing codes based on published studies^28–31^, and used Current Procedural Terminology (CPT) codes and International Classification of Diseases, 10th Revision, Procedure Coding System (ICD-10 PCS) codes for stroke imaging modalities in consultation with an expert vascular neurologist (coauthor A.S.). These billing codes and imaging codes are detailed in Supplemental Table 1. We identified 1,748 patients (1,288 ischemic strokes and 460 hemorrhagic strokes based on ICD codes) who had both an ICD-10 billing code as well as ICD-10 PCS or CPT code for cranial imaging during hospitalization.

We evaluated the performance of the billing/diagnosis plus imaging codes by selecting a random sample of 301 patients from the 1,748 identified (Supplemental Figure 1); two physicians conducted a detailed manual chart review for each hospitalization to determine whether each event was a stroke event or not. We defined a true-positive stroke as a patient with an *ICD-10* code and cranial imaging for stroke during index hospitalization, which was consistent with an adjudicated stroke event. We defined a false-positive stroke as a patient with a stroke code and imaging when no adjudicated stroke event was documented during hospitalization. The positive predictive value for the code-based approach was 80.4%.

To improve our ability to identify stroke, we augmented this using an artificial intelligence (AI) approach (Supplemental Figure 1). We used ChartScope, an AI-assisted chart review platform, to develop a scalable EHR machine learning/large language pipeline to identify acute incident stroke events and then extract stroke subtype (ischemic vs hemorrhagic). ChartScope was developed by coauthors R.C. and N.T. at Cedars-Sinai. The platform uses large language model–enabled querying of unstructured clinical notes using prompts to identify relevant patients more accurately and efficiently. Using sequential prompt engineering in the 301 patients who had undergone gold-standard physician adjudication, we developed a ChartScope prompt (Supplemental Methods) that achieved 89% accuracy, 93% PPV, 67% NPV, 95% sensitivity, and 63% specificity. Ischemic stroke was defined as any patient with clear evidence of an acute or subacute infarct, a patient who received intravenous thrombolysis for stroke or mechanical thrombectomy, or who had neurologist documentation of ischemic stroke. Hemorrhagic stroke was defined as any patient with either an intracerebral hemorrhage or subarachnoid hemorrhage, evidence of hemorrhagic conversion of an ischemic stroke, or neurologist documentation of hemorrhagic stroke. We found that ChartScope was 92% accurate in classification of type of stroke when compared to manual review, with most of the errors coming in patients who experienced hemorrhagic conversion of ischemic stroke. We then applied this prompt to all 1,748 code-based algorithmically defined patients (including the 301 adjudicated patients), yielding 1,344 LLM-defined stroke hospitalizations that formed the analytic cohort.

### Determination of mortality status, sudden cardiac arrest and cause of death determination

The 1,344 patients identified by the approach described above were followed until June 30, 2025, or until death. Stroke decedents (patients who died in hospital or discharged to hospice) were determined by using the EHR’s discharge status designation and confirming with death certificates. Stroke survivors were defined as patients who survived initial stroke hospitalization and were discharged successfully to home, to a post-acute care facility, or left against medical advice. Sudden cardiac deaths were determined from death certificate records from the California Department of Public Health.

We identified cases of presumed out-of-hospital cardiac arrest from the regional emergency medical services (EMS) database. Thereafter, trained physicians conducted a careful case adjudication using a standardized method as well as all available data, including medical records, EMS reports, and death certificates^26^. For this analysis, sudden cardiac arrest (SCA) was defined as a sudden and unexpected loss of pulse due to a likely cardiac cause^32^; patients whose cardiac arrest occurred due to trauma, substance use, or natural non-cardiac causes were excluded. Survivors of SCA (n=1, 1.5% of 67 total SCA cases) were included in the SCA outcome definition.

Cause of death was determined using the primary ICD-10 mortality code on the death certificate. These causes of death were then sorted into 10 categories: stroke/cerebrovascular, cardiac, cancer, other neurological causes (primarily Alzheimer’s Disease and other dementias), pulmonary, renal, infection/sepsis, diabetes/metabolic, GI/hepatic and other/unknown cases based on the leading causes of death as defined by the National Center for Health Statistics and other prior epidemiological papers^33,34^. Adjudicated SCA cases were treated as a subset of cardiac cause of death; all SCA cases except 1 were fatal.

### Statistical Analysis

Chi-square, Fisher’s exact tests (for cells with expected counts of ≤5), and Welch’s t-tests with two-sided *P* values <0.05 were used to evaluate statistically significant differences. For differences in survival analysis, the log-rank test was used. All statistical analysis and survival analyses were performed in Python [version 3.14] using the *lifelines* package [version 0.30.3] and Matplotlib [version 3.11.1], and two-sided *P* values <0.05 were considered statistically significant.

Age was analyzed as a continuous variable, reported as mean ± SD and median (IQR), and was also grouped into prespecified categories (<50, 50–74, 75–84, and ≥85 years) to characterize the age distribution of the cohort. Body-mass index was categorized according to World Health Organization criteria as underweight (<18.5 kg/m²), normal weight (18.5–24.9 kg/m²), overweight (25.0–29.9 kg/m²), obese class I (30.0–34.9 kg/m²), and obese class II or higher (≥35.0 kg/m²). BMI category was missing for 40 patients (3.0%) and was the only imputed variable; patients with an ICD-10 diagnosis code for obesity were assigned to obese class I, and those without were assigned to the normal-weight category for analysis.

Survival curves were generated for the overall cohort and stratified by stroke subtype (ischemic vs. hemorrhagic) and discharge disposition.

## RESULTS

### Study Population

The study population consisted of a total of 1,344 patients from the overall OSCAR cohort hospitalized with acute ischemic or hemorrhagic stroke (between January 1, 2017 and June 30, 2020) and followed through June 30, 2025. The mean age of this cohort was 74.0 ± 14.9 years, 629 patients (46.8%) were male, and 1,036 (77.1%) had ischemic and 308 (22.9%) hemorrhagic stroke. On the basis of stroke index-admission disposition, 1,141 patients (84.9%) survived to discharge (to home, a post-acute care facility, or against medical advice [AMA]), whom we designated as stroke survivors. The remaining 203 (15.1%) patients died during the index admission or were discharged to hospice (designated as stroke decedents). Over follow-up until June 30, 2025 (median follow-up 5.2 years, range 4 days to 8.5 years), 625 (54.8%) of the 1,141 stroke survivors died during the follow-up period.

### Baseline Characteristics: Stroke Decedents Versus Survivors (Table 1)

Stroke decedents and stroke survivors did not differ in age (75.2 ± 15.6 vs 73.8 ± 14.8 years; *P*=0.252) or male sex (42.4% vs 47.6%; *P*=0.169). However, stroke subtype differed significantly between groups: hemorrhagic stroke accounted for 46.3% of decedents but only 18.8% of survivors (*P*<0.001), and history of prior hemorrhagic stroke was more frequent among decedents (25.1% vs 14.3%; *P*<0.001). Decedents also carried a higher burden of prior cancer (45.8% vs 37.0%; *P*=0.017), whereas survivors more often had a prior transient ischemic attack (TIA; 42.6% vs 27.6%; *P*<0.001), hyperlipidemia (69.1% vs 59.1%; *P*=0.005), and a history of smoking (35.6% vs 25.6%; *P*=0.006). Body-mass index distribution also differed (*P*=0.030): survivors were more frequently overweight (36.4% vs 28.1%), whereas decedents were more frequently underweight (6.4% vs 3.2%). Conventional vascular risk factors and comorbidities (such as hypertension, type 2 diabetes, coronary artery disease, heart failure, prior myocardial infarction, and chronic kidney disease) did not differ significantly between groups.

**Table 1.** Baseline Characteristics: Stroke Survivors vs Stroke Deaths.

| **Variable** | Total<br>(n=1,344) | Stroke Survivors<br>(n=1,141) | Stroke Deaths<br>(n=203) | *P* value |
|---|---|---|---|---|
| **DEMOGRAPHICS** |  |  |  |  |
| **Male sex** | 629 (46.8%) | 543 (47.6%) | 86 (42.4%) | 0.169 |
| **Age, years** | 74.0 ± 14.9 | 73.8 ± 14.8 | 75.2 ± 15.6 | 0.252 |
| **Median (IQR)** | 76.2 (65.7-85.2) | 75.9 (65.4-84.9) | 77.9 (66.5-86.6) |  |
| **Age Category** |  |  |  | *P*=0.227 |
| **<50** | 93 (6.9%) | 79 (6.9%) | 14 (6.9%) |  |
| **50-74** | 539 (40.1%) | 468 (41.0%) | 71 (35.0%) |  |
| **75-84** | 368 (27.4%) | 313 (27.4%) | 55 (27.1%) |  |
| **85+** | 344 (25.6%) | 281 (24.6%) | 63 (31.0%) |  |
| **Race/Ethnicity** |  |  |  | *P*=0.017 |
| **White** | 816 (60.7%) | 690 (60.5%) | 126 (62.1%) |  |
| **Black** | 272 (20.2%) | 240 (21.0%) | 32 (15.8%) |  |
| **Hispanic** | 98 (7.3%) | 77 (6.7%) | 21 (10.3%) |  |
| **Asian** | 110 (8.2%) | 88 (7.7%) | 22 (10.8%) |  |
| **Other/Unknown** | 48 (3.6%) | 46 (4.0%) | 2 (1.0%) |  |
| **BMI Category** |  |  |  | *P*=0.030 |
| **Underweight** | 49 (3.6%) | 36 (3.2%) | 13 (6.4%) |  |
| **Normal Weight** | 518 (38.5%) | 428 (37.5%) | 90 (44.3%) |  |
| **Overweight** | 472 (35.1%) | 415 (36.4%) | 57 (28.1%) |  |
| **Obese Class I** | 198 (14.7%) | 170 (14.9%) | 28 (13.8%) |  |
| **Obese Class II+** | 107 (8.0%) | 92 (8.1%) | 15 (7.4%) |  |
| **Stroke Type** |  |  |  | *P*<0.001 |
| **Ischemic** | 1036 (77.1%) | 927 (81.2%) | 109 (53.7%) |  |
| **Hemorrhagic** | 308 (22.9%) | 214 (18.8%) | 94 (46.3%) |  |
| **COMORBIDITIES** |  |  |  |  |
| **Hypertension** | 1061 (78.9%) | 904 (79.2%) | 157 (77.3%) | 0.543 |
| **Type 2 Diabetes** | 538 (40.0%) | 459 (40.2%) | 79 (38.9%) | 0.725 |
| **Cancer Prevalence** | 515 (38.3%) | 422 (37.0%) | 93 (45.8%) | 0.017 |
| **Hyperlipidemia** | 909 (67.6%) | 789 (69.1%) | 120 (59.1%) | 0.005 |
| **Atrial Fibrillation** | 450 (33.5%) | 373 (32.7%) | 77 (37.9%) | 0.145 |
| **Coronary Artery Disease** | 644 (47.9%) | 551 (48.3%) | 93 (45.8%) | 0.515 |
| **CHF (All)** | 493 (36.7%) | 412 (36.1%) | 81 (39.9%) | 0.302 |
| **HFrEF** | 233 (17.3%) | 192 (16.8%) | 41 (20.2%) | 0.243 |
| **Prior MI** | 301 (22.4%) | 256 (22.4%) | 45 (22.2%) | 0.933 |
| **CKD** | 475 (35.3%) | 397 (34.8%) | 78 (38.4%) | 0.319 |
| **Valve Disease** | 527 (39.2%) | 444 (38.9%) | 83 (40.9%) | 0.596 |
| **Prior Stroke (Any)** | 691 (51.4%) | 585 (51.3%) | 106 (52.2%) | 0.804 |
| **Prior Ischemic Stroke** | 630 (46.9%) | 544 (47.7%) | 86 (42.4%) | 0.162 |
| **Prior TIA** | 542 (40.3%) | 486 (42.6%) | 56 (27.6%) | <0.001 |
| **Prior Hemorrhagic Stroke** | 214 (15.9%) | 163 (14.3%) | 51 (25.1%) | <0.001 |
| **Seizure** | 198 (14.7%) | 160 (14.0%) | 38 (18.7%) | 0.082 |
| **Smoking** | 458 (34.1%) | 406 (35.6%) | 52 (25.6%) | 0.006 |
| **Obesity** | 305 (22.8%) | 262 (23.0%) | 44 (21.7%) | 0.687 |
| **OUTCOMES** |  |  |  |  |
| **All-cause mortality** | 828 (61.6%) | 625 (54.8%) | 203 (100.0%) | <0.001 |
| **Sudden Cardiac Arrest** | 67 (5.0%) | 65 (5.7%) | 2 (1.0%) | 0.004 |

**Data are presented as No. (%) for categorical variables and mean ± SD for continuous variables, unless otherwise noted.**
**Stroke survivors were discharged home, to a post-acute care facility, or against medical advice; stroke deaths comprise in-hospital death and hospice discharge.**
***P* values were calculated using Welch's t-test for continuous variables and Pearson's chi-squared test for categorical variables, as indicated in the Test column.**
**AMA indicates discharge against medical advice; BMI, body mass index; CHF, congestive heart failure; CKD, chronic kidney disease; HFrEF, heart failure with reduced ejection fraction; IQR, interquartile range; TIA, transient ischemic attack.**

### Baseline Characteristics among post-discharge stroke Survivors, by Subtype (Table 2)

Among the 1,141 post-discharge survivors, 927 (81.2%) had ischemic and 214 (18.8%) had hemorrhagic stroke. Survivors of ischemic stroke were older than survivors of hemorrhagic stroke (74.9 ± 14.2 vs 68.9 ± 16.0 years; *P*<0.001) and had a higher prevalence of atherosclerotic and cardiac comorbidities, including hypertension (81.7% vs 68.7%; *P*<0.001), hyperlipidemia (70.9% vs 61.7%; *P*=0.009), coronary artery disease (50.1% vs 40.7%; *P*=0.013), prior myocardial infarction (23.6% vs 17.3%; *P*=0.045), and heart failure (37.6% vs 29.4%; *P*=0.024). Ischemic stroke survivors were more likely to have a history of a prior ischemic stroke (50.3% vs 36.4%; *P*<0.001) and prior TIA (45.4% vs 30.4%; *P*<0.001), whereas hemorrhagic-stroke survivors more frequently had a prior hemorrhagic stroke (40.7% vs 8.2%; *P*<0.001) and seizure history (22.4% vs 12.1%; *P*<0.001). Sex, race/ethnicity, body-mass index category, and discharge disposition did not differ by subtype.

**Table 2.** Baseline Characteristics of Stroke Survivors by Stroke Subtype.

| **Variable** | Ischemic | Hemorrhagic | *P* value |
|---|---|---|---|
|  | (n=927) | (n=214) |  |
| **DEMOGRAPHICS** |  |  |  |
| **Male sex** | 434 (46.8%) | 109 (50.9%) | 0.277 |
| **Age, years** | 74.9 ± 14.2 | 68.9 ± 16.0 | <0.001 |
| **Median (IQR)** | 76.9 (66.7-85.6) | 70.7 (58.2-80.3) |  |
| **Age Category** |  |  | *P*<0.001 |
| **<50** | 56 (6.0%) | 23 (10.7%) |  |
| **50-74** | 362 (39.1%) | 106 (49.5%) |  |
| **75-84** | 260 (28.0%) | 53 (24.8%) |  |
| **85+** | 249 (26.9%) | 32 (15.0%) |  |
| **Race/Ethnicity** |  |  | *P*=0.402 |
| **White** | 562 (60.6%) | 128 (59.8%) |  |
| **Black** | 199 (21.5%) | 41 (19.2%) |  |
| **Hispanic** | 59 (6.4%) | 18 (8.4%) |  |
| **Asian** | 67 (7.2%) | 21 (9.8%) |  |
| **Other/Unknown** | 40 (4.3%) | 6 (2.8%) |  |
| **BMI Category** |  |  | *P*=0.340 |
| **Underweight** | 29 (3.1%) | 7 (3.3%) |  |
| **Normal Weight** | 341 (36.8%) | 87 (40.7%) |  |
| **Overweight** | 338 (36.5%) | 77 (36.0%) |  |
| **Obese Class I** | 137 (14.8%) | 33 (15.4%) |  |
| **Obese Class II+** | 82 (8.8%) | 10 (4.7%) |  |
| **Location of Discharge** |  |  | *P*=0.458 |
| **AMA** | 6 (0.6%) | 2 (0.9%) |  |
| **Home** | 476 (51.3%) | 101 (47.2%) |  |
| **Post-acute care facility** | 445 (48.0%) | 111 (51.9%) |  |
| **COMORBIDITIES** |  |  |  |
| **Hypertension** | 757 (81.7%) | 147 (68.7%) | <0.001 |
| **Type 2 Diabetes** | 382 (41.2%) | 77 (36.0%) | 0.16 |
| **Cancer Prevalence** | 332 (35.8%) | 90 (42.1%) | 0.088 |
| **Hyperlipidemia** | 657 (70.9%) | 132 (61.7%) | 0.009 |
| **Atrial Fibrillation** | 315 (34.0%) | 58 (27.1%) | 0.053 |
| **Coronary Artery Disease** | 464 (50.1%) | 87 (40.7%) | 0.013 |
| **CHF (All)** | 349 (37.6%) | 63 (29.4%) | 0.024 |
| **HFrEF** | 158 (17.0%) | 34 (15.9%) | 0.684 |
| **Prior MI** | 219 (23.6%) | 37 (17.3%) | 0.045 |
| **CKD** | 333 (35.9%) | 64 (29.9%) | 0.096 |
| **Valve Disease** | 364 (39.3%) | 80 (37.4%) | 0.611 |
| **Prior Stroke (Any)** | 482 (52.0%) | 103 (48.1%) | 0.308 |
| **Prior Ischemic Stroke** | 466 (50.3%) | 78 (36.4%) | <0.001 |
| **Prior TIA** | 421 (45.4%) | 65 (30.4%) | <0.001 |
| **Prior Hemorrhagic Stroke** | 76 (8.2%) | 87 (40.7%) | <0.001 |
| **Seizure** | 112 (12.1%) | 48 (22.4%) | <0.001 |
| **Smoking** | 328 (35.4%) | 78 (36.4%) | 0.769 |
| **Obesity** | 219 (23.6%) | 43 (20.1%) | 0.268 |
| **OUTCOMES** |  |  |  |
| **All-cause mortality** | 512 (55.2%) | 113 (52.8%) | 0.52 |
| **Median survival** | 2026 d (5.5 yr) | 2107 d (5.8 yr) | 0.898 |
| **Sudden Cardiac Arrest** | 56 (6.0%) | 9 (4.2%) | 0.296 |

**Data are presented as No. (%) for categorical variables and mean ± SD for continuous variables, unless otherwise noted.**
**Stroke survivors were discharged home, to a post-acute care facility, or against medical advice.**
***P* values were calculated using Welch's t-test for continuous variables, Pearson's chi-squared test without continuity correction for categorical variables, and the Fisher-Freeman-Halton exact test for location of discharge (expected cell count ≤5). Median survival was estimated with the Kaplan-Meier method and compared with the log-rank test.**
**AMA indicates discharge against medical advice; BMI, body mass index; CHF, congestive heart failure; CKD, chronic kidney disease; HFrEF, heart failure with reduced ejection fraction; IQR, interquartile range; MI, myocardial infarction; TIA, transient ischemic attack.**

### Long term survival after discharge by stroke subtype and discharge location (Figures 1 and 2)

Median survival among patients who were discharged from the hospital after index stroke was 5.5 years (2,026 days) after ischemic stroke and 5.8 years (2,107 days) after hemorrhagic stroke, and all-cause mortality during follow-up did not differ significantly between subtypes (*P*=0.90, Figure 1).

**Figure 1.**
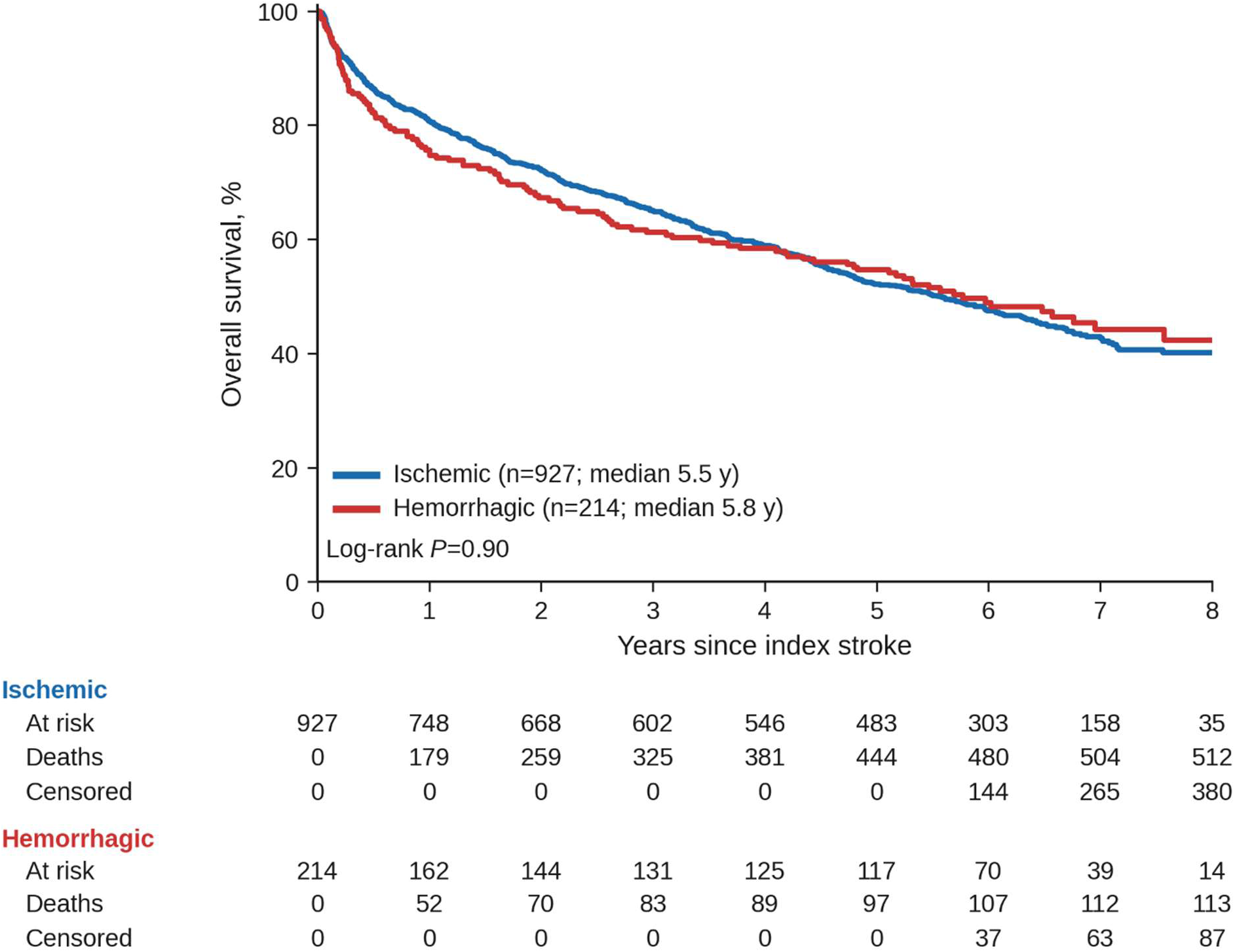
Kaplan-Meier Survival Curves by Stroke subtype among patients discharged from the hospital. Ischemic stroke median survival=5.5 years, hemorrhagic stroke median survival=5.8 years (χ²=0.003 by log-rank test, *P*=0.90).

**Figure 2.**
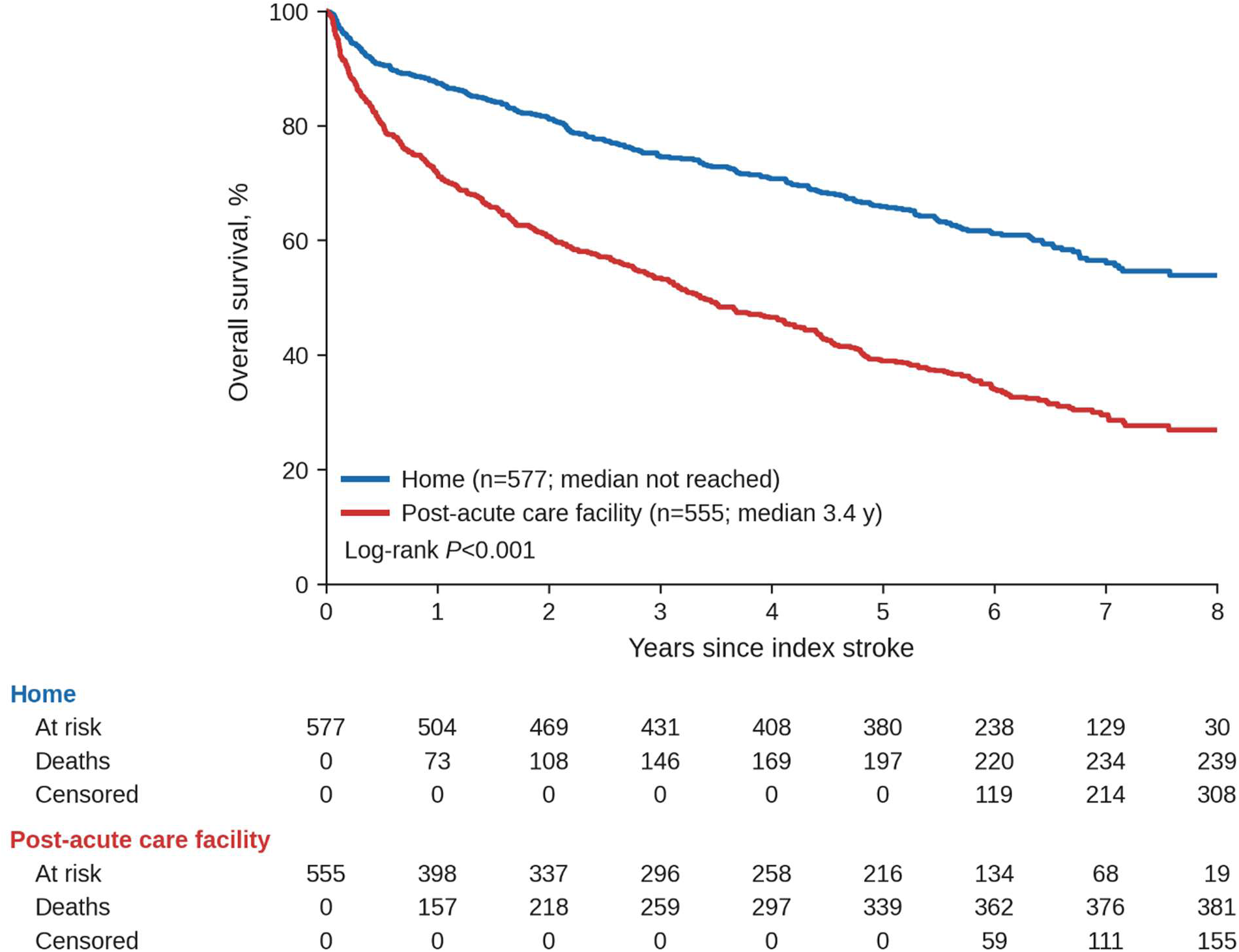
Kaplan-Meier Survival Curves by discharge location (home vs post-acute care facility). Post-acute care median=3.4 years. Home median not reached. (χ²=93.9, *P*<0.001 by log-rank test).

Median survival among patients discharged home was >8.5 years (more than half of these patients were alive at the end of follow-up, so exact median survival cannot be calculated), whereas median survival among patients discharged to a post-acute care facility was 3.4 years (1,226 days) (*P*<0.001, Figure 2).

### Causes of Death: Acute Decedents Versus Post-Discharge Deaths (Figure 3)

The distribution of physician-adjudicated causes of death differed significantly between the 203 acute stroke decedents and the 625 deaths occurring after discharge among survivors of the initial hospitalization for stroke (*P*<0.001). Cerebrovascular causes predominated among acute decedents, accounting for 36.0% of deaths versus 15.5% of post-discharge deaths (*P*<0.001). In contrast, cardiac causes were the leading mode of death after discharge, comprising 28.8% of post-discharge deaths versus 16.3% of acute deaths (*P*<0.001). Cancer accounted for a nearly identical proportion of deaths in both groups (approximately 22%). Other neurological (8.5% vs 3.4%; *P*<0.05) and infectious/septic (8.8% vs 3.4%; *P*<0.05) causes were also more frequent among post-discharge deaths than among stroke decedents.

**Figure 3.**
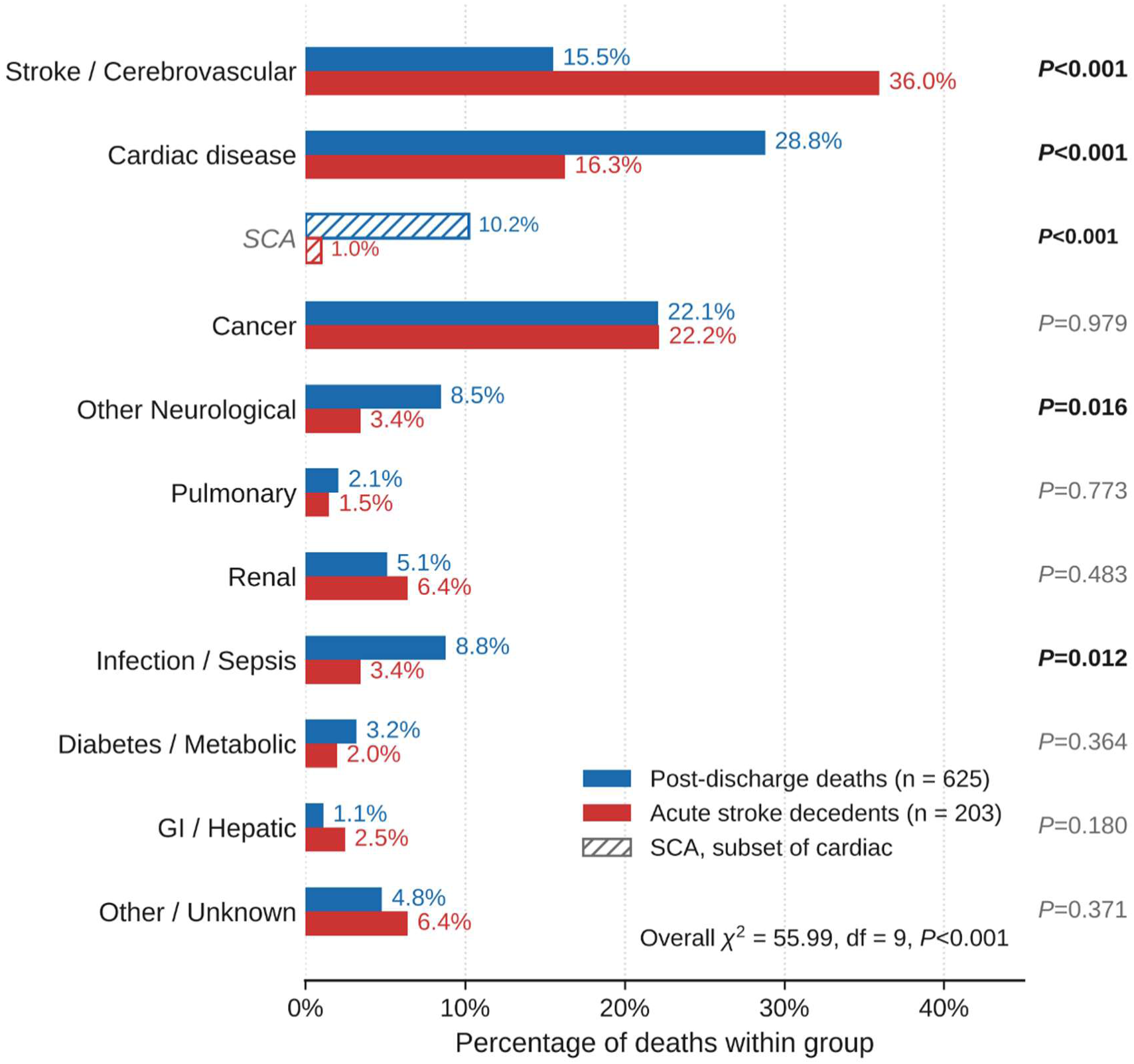
Adjudicated causes of death among post-discharge deaths among initial survivors (post-discharge deaths, n = 625) versus acute stroke decedents (n = 203). Bars represent the proportion of deaths within each group. Sudden cardiac arrest (SCA, hatched) is shown as a subset of cardiovascular causes. Overall χ²=55.99, df=9, *P*<0.001. Individual *P* values for each cause also shown.

### Causes of Death Among Survivors, by Stroke Subtype (Figure 4)

Among the 625 post-discharge deaths (512 after ischemic and 113 after hemorrhagic stroke), the cause-of-death distribution differed significantly by index stroke subtype (*P*<0.001). For ischemic stroke survivors, cardiac disease was the leading cause of death (31.6%), followed by cancer (18.4%) and stroke/cerebrovascular disease (16.4%). For hemorrhagic stroke survivors, the pattern was reversed: cancer was the leading cause (38.9%), followed by cardiac disease (15.9%) and stroke/cerebrovascular disease (11.5%). Among patients with no known history of cancer before stroke, cancer accounted for twice the proportion of post-discharge deaths after hemorrhagic stroke as after ischemic stroke (19.3% vs 9.6%; 11.3% among all stroke survivors).

**Figure 4.**
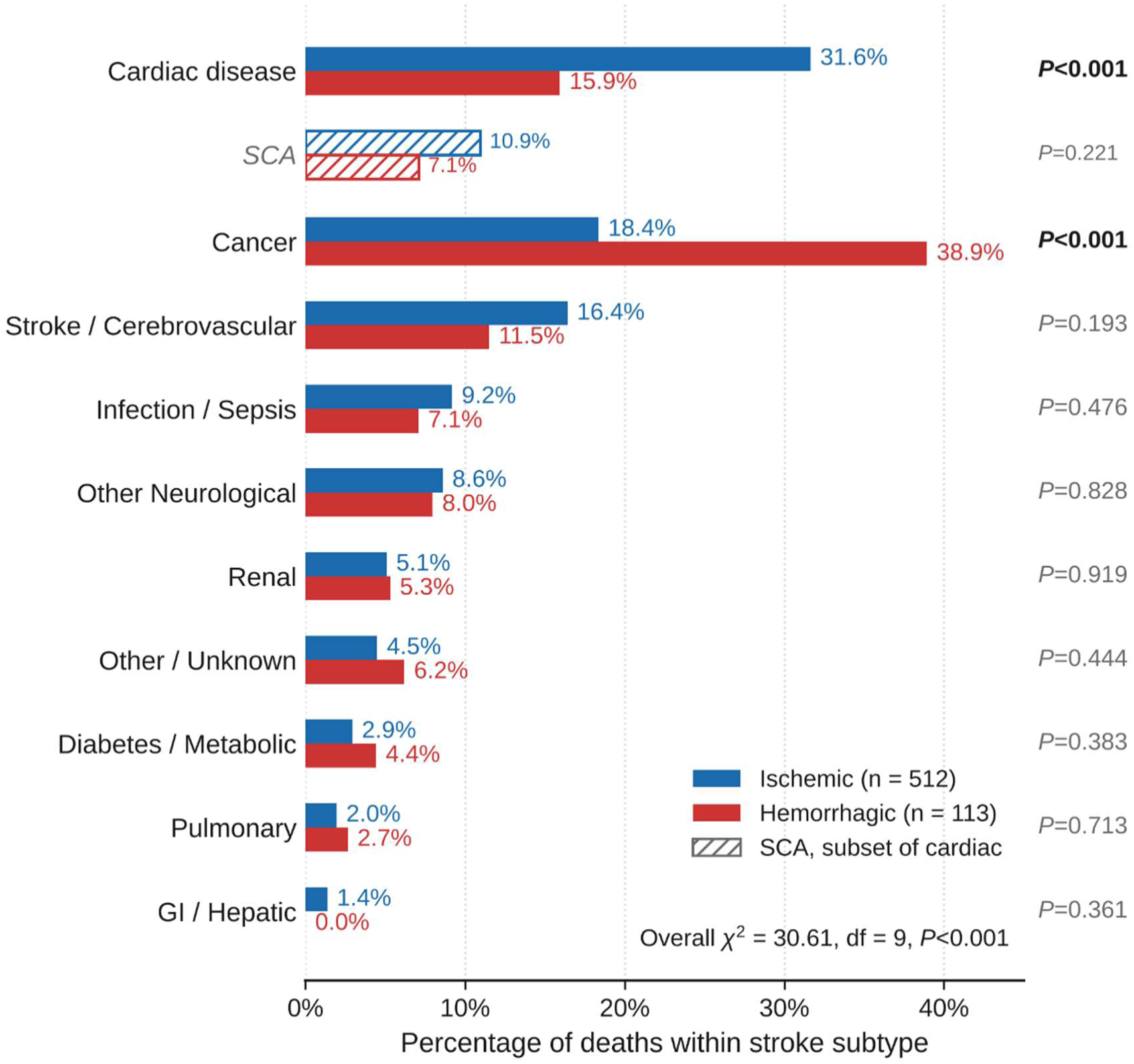
Adjudicated causes of death among discharged stroke survivors who died during follow-up, stratified by index stroke subtype (ischemic, n = 512; hemorrhagic, n = 113). SCA (hatched) is shown as a subset of cardiac disease. Overall χ²=30.61, df=9, *P*<0.001. Individual *P* values for each cause also shown.

Sudden cardiac arrest (SCA) was a prominent mode of death in both stroke subtypes, but ischemic-stroke survivors were disproportionally affected. As a clinical event during follow-up, SCA occurred in 56 ischemic-stroke survivors (6.0%) versus 9 hemorrhagic-stroke survivors (4.2%; *P*=0.296). The disparity was more pronounced among the subset of patients who died during the follow-up period. Among the 625 survivors who died during follow-up, SCA accounted for 10.9% of all deaths after ischemic stroke compared with 7.1% after hemorrhagic stroke. Overall, cardiac disease was responsible for 31.6% of deaths after ischemic stroke versus 15.9% after hemorrhagic stroke; SCA thus represented roughly one-third of all cardiac deaths in the ischemic group and nearly half in the hemorrhagic group.

## DISCUSSION

In this contemporary cohort of 1,344 patients hospitalized with stroke in which survivors were followed for a median of 5.2 years, causes of death shifted from cerebrovascular disease during the index hospitalization to cardiac disease after discharge and diverged sharply by stroke subtype. Overall mortality in our cohort was consistent with contemporary US estimates. Early mortality (in-hospital death or discharge to hospice) in our cohort was 10.5% after ischemic stroke and 30.5% after hemorrhagic stroke, compared with national population-based 30-day case fatality of 8.1% for ischemic stroke, 39.5% for intracerebral hemorrhage, and 17.5% for subarachnoid hemorrhage^35^. In our study, cerebrovascular causes accounted for 36.0% of deaths during the index hospitalization but only 15.5% of deaths after discharge, when cardiac disease became the leading cause (28.8%). This cardiac predominance was most evident after ischemic stroke, in which cardiac disease caused 31.6% of post-discharge deaths and SCA alone accounted for 10.9%, approximately one-third of cardiac deaths. After hemorrhagic stroke, cancer was the leading cause of post-discharge death (38.9%). Despite these divergent profiles, overall survival after discharge did not differ by subtype, indicating that similar all-cause mortality can conceal fundamentally different causes of death.

Much of the current understanding of mortality in stroke survivors is derived from population-based cohorts assembled before endovascular thrombectomy became widespread. In those cohorts, cerebrovascular and cardiac causes were often combined, with little data on the specific causes of cardiac mortality. In addition, those studies combined hemorrhagic and ischemic stroke and thus did not investigate whether there was a difference in mortality patterns based on stroke subtype^2–5,16^. Contemporary cause-of-death data, by contrast, are sparse and largely limited to the first weeks after the index event, when deaths are driven mainly by the index stroke and its complications^25^. Our data show that this temporal difference in cause of death persists in a contemporary cohort. The dominant late cause differs by subtype, with cardiac and oncologic contributions after ischemic and hemorrhagic stroke, respectively, that exceed what pooled historical series would predict. In prior studies, cancers were the cause of death in 7%–11.8% of patients, and cardiac causes of death occurred in 20%–23% of patients, but these studies largely pooled ischemic and hemorrhagic stroke^2–5^. Another strength of our study is the systematic, physician-adjudicated ascertainment of SCA assisted by EMS records.

The cardiac predominance after ischemic stroke is consistent with prior work and likely reflects overlapping processes^6,10^. First, ischemic stroke and coronary disease share an atherosclerotic substrate, and ischemic stroke survivors in our cohort carried a high burden of hypertension, hyperlipidemia, coronary artery disease, prior myocardial infarction, and heart failure. Second, stroke can itself injure the heart through autonomic dysregulation, catecholamine surge, and neurogenic myocardial damage, a constellation termed the stroke–heart syndrome^12^. Cardiac complications develop in 10% to 20% of patients within days of stroke and are independently associated with 90-day mortality^12,13^. Atrial fibrillation detected after stroke may partly reflect this neurogenic substrate^11,14^. Sudden cardiac death (SCD) is a recognized but frequently under-attributed consequence of these mechanisms^15,36^. However, secondary prevention after ischemic stroke may not include dedicated imaging to evaluate coronary perfusion; our study raises the notion that some segment of these stroke survivors may benefit from evaluation and treatment of undiagnosed ischemic heart disease and extended rhythm monitoring for SCA^37–40^.

The prominence of SCA is consistent with growing evidence that stroke is itself a risk factor for SCD, not merely a marker of shared vascular risk^15,41^. During continuous monitoring in the first 72 hours after acute ischemic stroke, serious ventricular arrhythmias are detected in approximately 5% of patients and atrial arrhythmias in approximately 20%^42^. Because atrial fibrillation is a recognized cause of ischemic stroke, its detection after the event does not establish whether it preceded or followed the stroke. In contrast, more research is needed to understand whether detection of ventricular arrhythmia post-stroke is secondary to pre-established cardiac disease and a pre-stroke risk factor, versus whether it develops post-stroke secondary to cardiac autonomic dysregulation triggered by the stroke^14^. Insular cortex injury has been linked to disturbed cardiac autonomic control and myocardial injury^43–45^. Among patients with a first ischemic stroke in the Northern Manhattan Study, sudden death accounted for the largest share of cardiac deaths, and left parietal infarction independently predicted cardiac death^6^. Post-stroke repolarization abnormalities, including QT prolongation, have likewise been linked to malignant dysrhythmias but remain under-recognized^36^. Together with our observation that SCA constituted one-third of cardiac deaths after ischemic stroke, these data support viewing ischemic stroke survivors as a population at risk for SCD, consistent with the stroke–heart syndrome framework^10,12^. Beyond monitoring for atrial fibrillation, structured longitudinal assessment of cardiac and sudden-death risk, including electrocardiographic abnormalities, ischemic evaluation, left ventricular function, and heart failure phenotype, may be warranted in the months and years after discharge, when the competing neurological hazard has receded. Identifying the stroke survivors at highest risk of SCD is an actionable target for a prospective study.

After hemorrhagic stroke, cancer accounted for more than twice the proportion of post-discharge deaths observed after ischemic stroke (38.9% vs 18.4%). Prior cancer was more common among acute decedents than survivors (45.8% vs 37.0%) and, among survivors, tended to be more frequent after hemorrhagic than ischemic stroke (42.1% vs 35.8%), raising the possibility that, in some patients, malignancy did not merely coexist with the hemorrhagic stroke but may have also contributed to it. Indeed, even among patients with no known history of cancer before stroke, cancer accounted for twice the proportion of post-discharge deaths after hemorrhagic stroke as after ischemic stroke (19.3% vs 9.6%; 11.3% among all stroke survivors). This is biologically plausible: in a large series of patients with cancer, intratumoral hemorrhage and coagulopathy accounted for most intracranial hemorrhages, whereas hypertension was implicated in approximately 5%^46^. Thrombocytopenia and treatment-related vascular injury provide additional pathways^46,47^. Historical descriptions of survivorship after ICH, which carries a high early case fatality, identify cerebrovascular disease as the leading long-term cause of death, although these deaths occur largely soon after the index event^7,8^. Among patients in our cohort who survived to discharge after hemorrhagic stroke, malignancy instead predominated. Thus, occult or active malignancy merits consideration in the evaluation of ICH without a clear hypertensive or amyloid explanation, consistent with guideline recommendations to investigate secondary causes^48^. Prognosis after hemorrhagic stroke may also depend more on cancer prognosis than on conventional vascular risk factors.

Collectively, these findings indicate that pooling of stroke subtypes obscures two clinically distinct mortality trajectories and that subtype-specific follow-up, with cardiac and dysrhythmic surveillance after ischemic stroke and oncologic evaluation after hemorrhagic stroke, may serve survivors better than a single generic post-stroke pathway. Discharge disposition was also strongly associated with long-term survival. Patients discharged home lived substantially longer than those discharged to a post-acute care facility: more than half of those discharged home were still alive at the end of follow-up, so their median survival could not be estimated, whereas median survival after discharge to a post-acute care facility was 3.4 years. The survival curves separated early, consistent with frailty and functional dependence after stroke as important determinants of long-term outcome.

### Limitations

This study has limitations. First, it was conducted in a single health system in Los Angeles; although the cohort was demographically diverse, the case mix of a quaternary academic center may limit generalizability. Second, stroke identification relied on a hybrid billing-code, imaging, and large language model–assisted pipeline; despite a high positive predictive value (92.9%) and sensitivity (95%) against physician adjudication, its lower specificity (63.3%) and negative predictive value (67.4%) imply some misclassification of stroke status. Third, individual-level data on reperfusion therapy, stroke severity, medications, lesion characteristics, and functional status were not available, so we could not examine whether these factors modified cause-specific mortality. Fourth, except for adjudicated SCA, cause of death was based on the primary ICD-10 code on the death certificate, which is subject to misclassification^49^.

## Conclusions

In this cohort, the leading cause of death after discharge differed by stroke subtype: cardiac disease, including sudden cardiac arrest, after ischemic stroke and cancer after hemorrhagic stroke. These findings support subtype-specific post-stroke follow-up and warrant further prospective evaluation of structured cardiac and sudden-death risk assessment in ischemic stroke survivors.

## Ethical statement

The OSCAR study was approved by the institutional review board (IRB) at CSHS and by the Honest Enterprise Research Broker (HERB) Committee of CSHS’s Enterprise Information Systems (EIS) Research Informatics and Scientific Computing Core (RISCC).

## Data Availability

Deidentified participant data will be made available after publication upon reasonable request to the corresponding author, following approval of a proposal and a signed data use agreement.

## Supplemental Material

Supplemental Methods

Figure S1

Table S1

## Nonstandard Abbreviations and Acronyms

AMA: against medical advice
EHR: electronic health record
EMS: emergency medical services
ICH: intracerebral hemorrhage
LLM: large language model
NPV: negative predictive value
SCA: sudden cardiac arrest
OSCAR: Observational Study of Cardiac Arrest Risk
PPV: positive predictive value
SCD: sudden cardiac death
TIA: transient ischemic attack

